# A Cerebral Frailty Risk Score Integrating Frailty Index and Neuroimaging for Dementia Prediction in the UK Biobank

**DOI:** 10.64898/2026.04.01.26350015

**Authors:** Cheuk Ni Kan, Justin Chew, Wee Shiong Lim, Chin Hong Tan

**Affiliations:** Psychology, School of Social Sciences, Nanyang Technological University, Singapore; Institute of Geriatrics and Active Aging, Tan Tock Seng Hospital, Singapore; Department of Geriatric Medicine, Tan Tock Seng Hospital, Singapore; Lee Kong Chian School of Medicine, Nanyang Technological University, Singapore

## Abstract

Frailty is a multisystem clinical syndrome closely linked to cognitive aging, yet its cerebral underpinnings and co-contribution to adverse outcomes remain poorly understood. In 63,509 dementia-free UK Biobank participants (aged 65.0±7.7), higher frailty index (FI) was associated with multiple neuroimaging markers, including reduced hippocampal volume, decreased cortical thickness, greater white matter hyperintensities burden, and impaired brain diffusion metrics. FI and neuroimaging markers additively increased the risks of incident dementia and mortality. An extreme gradient boosting with accelerated failure time framework highlighted FI and key regional neuroimaging features in dementia risk prediction (nested C-index=0.825, iAUC=0.759). Integrating the top 10 predictors into a novel point-based cerebral frailty risk score (CFRS) showed strong performance in predicting dementia onset (optimism-corrected C-index=0.838, iAUC=0.778), and was robust to the competing risk of mortality. These findings highlight the potential utility of a CFRS framework that integrates cumulative systemic and cerebral vulnerabilities for dementia risk stratification.

## INTRODUCTION

Frailty in older adults is a major public health challenge in aging populations^1^ and is broadly described as a state of deterioration in multiple physiological systems that increases vulnerability to acute stressors^2^. Although closely related to chronological age, frailty represents a distinct clinical syndrome associated with a range of adverse health outcomes, including increased risk of falls, functional disability, hospitalization, mortality^2–5^, incident stroke, and poorer stroke recovery^6^. Frailty has traditionally been characterized using the Fried phenotype, which emphasizes physical indicators such as slow gait speed and muscular weakness^2^. However, it is also commonly conceptualized using a deficit accumulation approach, which captures the multidimensional burden of clinical symptoms and comorbidities reflecting individual variations in the aging process^5,7,8^.

The close relationship between frailty, cognitive function, and risk of incident neurocognitive disorders has also been recognized^9–11^, leading to the conceptualization of “cognitive frailty”^12^. Just as frailty represents the cumulative burden of multisystem physiological decline, cognitive impairment reflects cerebral dysfunction arising from a range of underlying neuropathological changes. Emerging evidence suggests that structural brain health may partially account for the association between physical frailty and poor health outcomes^13^, and may also buffer mortality risk among frail individuals^14^. These findings point to shared biological substrates linking brain and physical health and underscore the central role of cerebral integrity in shaping frailty-health trajectories. Hence, there is a critical need to elucidate the neural underpinnings of frailty to gain mechanistic insights into how cerebral dysfunction in conjunction with frailty contributes to adverse health outcomes such as dementia, stroke, and mortality.

To date, only a limited number of studies have investigated the relationship between frailty and selected measures of white matter health^13,15–18^ and cerebral volumetrics^13,16,18–20^, with inconsistent results. Most prior investigations were also constrained by modest sample sizes and focused predominantly on the physical phenotype of frailty, thereby understating its multidimensional nature. The restricted scope of neuroimaging measures has further hindered a more comprehensive neural characterization of frailty and identification of key predictors of adverse outcomes. While increasing evidence suggests that machine learning approaches can outperform traditional proportional hazards models in disease predictions by flexibly analyzing diverse and large volumes of biomedical data^21–23^, common techniques such as tree-based models may also have notable drawbacks. These include reduced interpretability and inadequate handling of observation windows and censoring. Consequently, analytical approaches that explicitly model time-to-event while accounting for censoring may be better suited for capturing slow-progressing conditions such as dementia^24^ and the accumulative nature of frailty and cerebral dysfunction over time.

To enable a more holistic evaluation of the neural correlates of frailty and their contributions to major health outcomes, this study leveraged large-scale, population-based clinical and neuroimaging data, and applied a combination of machine learning approaches to translate complex survival models into an interpretable clinical risk score. Specifically, we sought to: (1) characterize neuroimaging markers associated with frailty based on the deficit accumulation approach^5^; (2) determine how frailty and cerebral dysfunction jointly contribute to the risks of incident dementia, stroke, and mortality; and (3) integrate frailty and multimodal neuroimaging to develop a novel cerebral frailty risk score (CFRS) for dementia prediction. We hypothesized that the multisystem nature of frailty would be reflected in associations with multiple indices of brain aging, and that their joint consideration within a CFRS framework would best predict dementia risk.

## METHODS

### Participants

The UK Biobank (http://www.ukbiobank.ac.uk/) is a multicenter prospective cohort study of over 500,000 individuals aged 40 to 69 years, recruited across the United Kingdom between 2006 and 2010^25^. All participants underwent baseline assessments, including biological sample collection for laboratory analysis, physical examination, and an extensive questionnaire covering health conditions, lifestyle factors, and medical history. The UK Biobank database also contains information on participants’ subsequent health outcomes via linkage to the UK National Health Service records. Since 2014, over 100,000 participants have been invited back for multimodal imaging and repeated questionnaire assessments (i.e. the imaging visit), which were primarily used in this study (Figure 1). Ethics approvals for the UK Biobank were obtained from the Multi-Centre Research Ethics Committee and the Human Tissue Authority for research tissue banking^25^.

**Figure 1.**
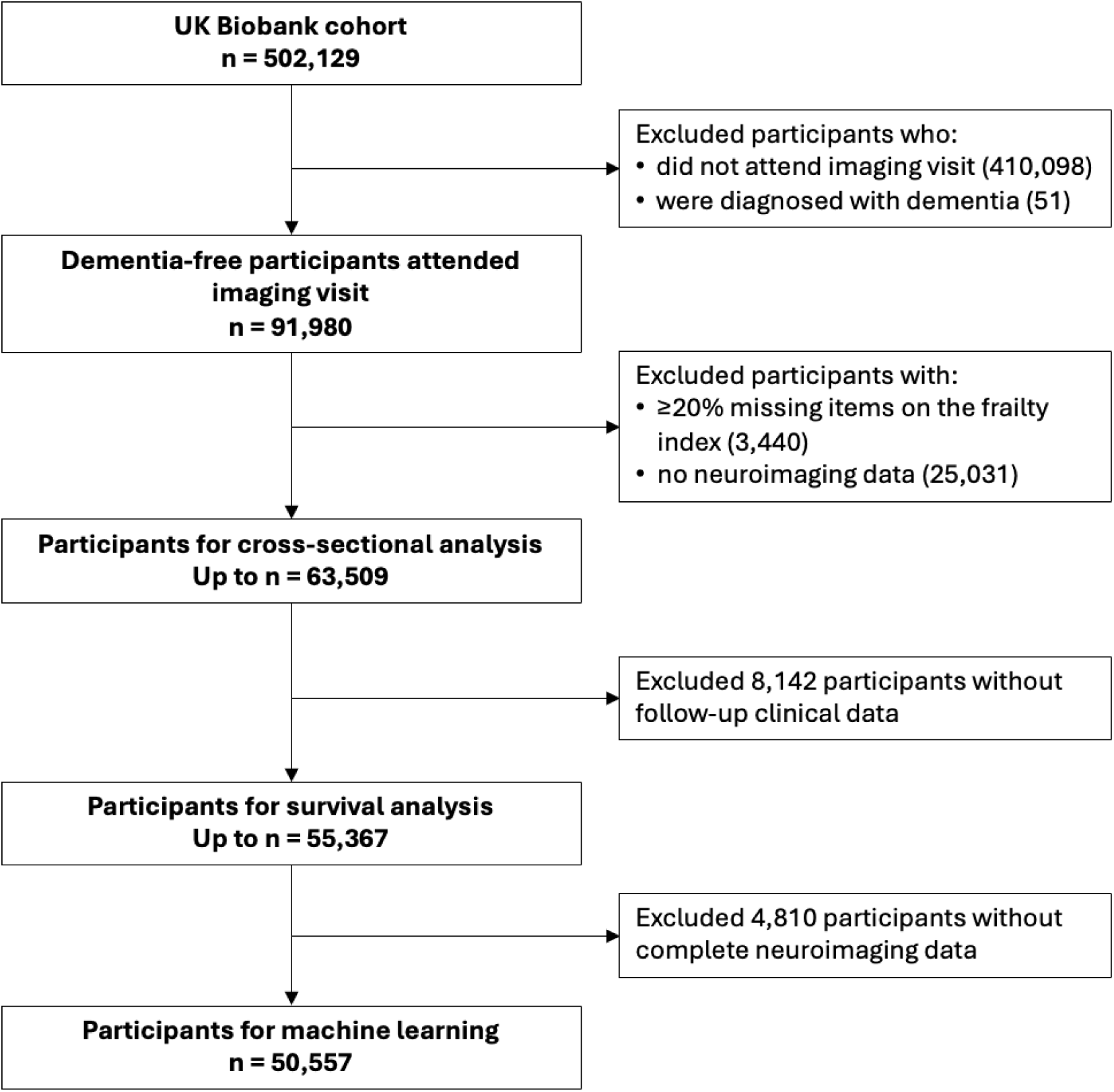
Sample selection. Dementia-free participants from the UK Biobank cohort were selected based on the availability of clinical and neuroimaging data.

### Clinical Outcomes

This study investigated incident dementia as the primary outcome, and incident stroke and mortality as the secondary outcomes. All-cause dementia and stroke diagnoses were ascertained using the UK Biobank’s algorithmically-defined outcomes^26^ that combined hospital records, mortality data, primary care data, and self-report, coded according to the International Statistical Classification of Diseases, 10^th^ Revision (ICD-10). Participants with prevalent dementia were excluded from the cohort, and those whose stroke dates preceded their imaging visit dates were subsequently excluded from the analyses of time to incident stroke. Mortality status and date of death within the UK Biobank database were determined through linkage to national death registries.

### Frailty Index

Frailty was operationalized as a frailty index (FI) that was previously constructed and validated within the UK Biobank cohort^5^. The FI was derived from 49 symptoms, diagnosed diseases, and functional impairments across multiple physiological and mental domains, including sensory (i.e. eye conditions, hearing difficulty), cranial, mental well-being (e.g. mood disturbances, loneliness), infirmity (i.e. falls, fractures, disability), cardiometabolic (e.g. diabetes, hypertension, hyperlipidemia), respiratory, musculoskeletal, immunological, cancer, pain, and gastrointestinal. Each variable was coded between 0 (no deficit) and 1 (most severe deficit), and individual FI values were derived following the deficit accumulation approach^7^ by dividing the sum of accrued deficits by the total number of measured deficits in each participant. All FI variables and their coding have been previously documented^5^. Consistent with previous protocol^5^, participants who had missing data for 10 or more (>20%) variables for FI derivation were excluded from the sample.

### Neuroimaging and Derived Phenotypes

All brain magnetic resonance imaging (MRI) scans in the UK Biobank cohort were performed on identical 3T Siemens Skyra scanners with standard Siemens 32-channel head coil. Details of the acquisition parameters, preprocessing, and quality control procedures have been extensively described^27,28^. We extracted neuroimaging-derived global and regional phenotypes that were generated by the UK Biobank image processing pipeline. Brain volumetrics were acquired using structural T1-weighted sequences (voxel size = 1.0 × 1.0 × 1.0 mm^3^, matrix size = 208 × 256 × 256) and processed with FreeSurfer 6.0 automatic segmentation for subcortical volumes and cortical parcellation based on the Desikan-Killiany atlas^29^ for regional cortical thickness. White matter hyperintensities (WMH) volumes were derived from the T2-weighted fluid-attenuated inversion recovery sequences (voxel size = 1.05 × 1.0 × 1.0 mm^3^, matrix size = 192 × 256 × 256) and processed with Brain Intensity AbNormality Classification Algorithm (BIANCA)^30^ for automated lesion segmentation. We performed log-transformation on the WMH volumes to adjust for skewness. Microstructural measures were derived from diffusion MRI (voxel size = 2.0 × 2.0 × 2.0 mm^3^, matrix size = 104 × 104 × 72) fitted with diffusion tensor imaging (DTI, b = 1000 s/mm² shell) and processed with Tract-Based Spatial Statistics (TBSS)^31^ for tract-wise fractional anisotropy (FA) and mean diffusivity (MD). Brain perfusion data were acquired using arterial spin labeling (ASL) sequences (voxel size = 3.4 × 3.4 × 4.5 mm^3^, matrix size = 64 × 48 × 32) with multi-postlabeling delay (PLD) pCASL acquisition (PLDs = 400/800/1200/1600/2000 ms) and processed with FSL’s Bayesian Inference for ASL MRI (BASIL) toolbox^32^ for cerebral blood flow (CBF) quantification. As ASL MRI was added to the UK Biobank imaging protocol in February 2021, only a subset of the cohort had CBF data. For each region-of-interest, we averaged bilateral measures of cortical thickness, FA, MD, and CBF and summed bilateral subcortical volumes prior to statistical analysis to reduce noise and multiple comparisons.

### Statistical Analysis

We conducted multiple linear regression to examine whether global neuroimaging measures of neurodegeneration (global cortical thickness, hippocampal volume), cerebrovascular health (total WMH volume, global CBF), and microstructural integrity (global FA and MD) were associated with higher FI values in separate models, adjusted for age at imaging visit, sex, and years of education. Total intracranial volume (TIV) was additionally included as a covariate in models involving structural volumetrics (hippocampus and WMH) to adjust for head size. Prior to analysis, education qualifications in the UK Biobank were converted to years of education^33^, and the global neuroimaging measures and FI values were scaled to provide standardized estimates.

We conducted Cox proportional hazards regression to examine the association of FI values and global neuroimaging measures with risks of incident all-cause dementia, stroke, and mortality, adjusted for age, sex, and years of education, as well as TIV in models with structural volumetrics. Several models were fitted: 1) FI as the main predictor, 2) FI and each global neuroimaging measure as the main predictors, 3) interaction between FI and each neuroimaging measure with their based terms. For any statistically significant interaction, a simple slopes analysis was conducted to determine how the effect of FI was modified by the neuroimaging marker. Global tests of scaled Schoenfeld residuals confirmed the proportional hazards assumption for all models predicting dementia and incident stroke (*p* > 0.05). As age showed time-varying effect in the mortality models, we stratified by age group to address the non-proportionality. CBF measures were excluded from this analysis as only a small subset of the cohort had CBF data. When separate models were fitted for each neuroimaging marker, the *p*-values were corrected for multiple comparisons using the false discovery rate (FDR) approach. All analyses were conducted using R 4.4.2.

### Machine Learning Approach

#### Candidate predictors

In participants with complete clinical and neuroimaging data, we implemented an extreme gradient boosting-based accelerated failure time (XGBoost-AFT)^34^ model (*xgboost* package) to predict time to incident all-cause dementia while accounting for right censoring. The cerebral frailty model comprised of demographic variables (age, sex, years of education), FI values, neuroimaging phenotypes, and TIV. A total of 130 neuroimaging variables were used, including regional cortical thickness measures, cortical and subcortical volumetric measures, tract-level FA and MD values, and WMH volumetric measures. An overview of all variables used for model building is provided in Supplementary Table S1. All continuous variables were scaled, and the sex variable was one-hot encoded prior to model fitting.

#### Model training and tuning

Given the low number of events (n = 106), model development and evaluation were performed using a nested cross-validation (CV) framework to obtain approximately unbiased performance estimates^35^. The outer loop used 5-fold stratified CV based on the event indicator to create independent test folds for estimating the generalization accuracy. Within each outer training fold, hyperparameters were tuned via 3-fold inner CV using Bayesian optimization (*ParBayesianOptimization* package), which maximized the mean validation AFT negative log-likelihood across inner folds. The tuning search space included maximum tree depth, learning rate, row subsampling, column subsampling, minimum child weight, and regularization terms (gamma, L1 and L2). The final model was trained with the optimal hyperparameter set on each outer training fold and evaluated on the corresponding outer test fold. Inverse-probability sample weights based on event frequency were incorporated during training to adjust for event imbalance and improve model stability^36^. Model predictions from all outer test folds were pooled to compute performance metrics.

#### Model evaluation and interpretation

Model discrimination was evaluated using the concordance index (C-index)^37^ and cumulative/dynamic time-dependent area under the receiver operating characteristic curve (AUC)^38^ across time horizons of 1 to 7 years and integrated AUC (*timeROC* package). Overall prediction accuracy was quantified using the integrated inverse-probability-of-censoring-weighted Brier scores (iBS)^39^. For model interpretability, Shapley Additive exPlanations (SHAP)^40^ values were computed for each outer test fold (*SHAPforxgboost* package) and pooled to derive unbiased global feature importance based on mean absolute SHAP magnitude. We plotted the SHAP summary to identify the top 10 contributing predictors for downstream development of an integer-based risk score.

#### Cerebral frailty risk score derivation and evaluation

We implemented AutoScore-Survival^41^ (*AutoScore* package), a machine learning-based clinical score generator, to construct an interpretable CFRS for dementia using the top SHAP-ranked variables. The AutoScore-Survival workflow performed automated variable discretization and quantile-based categorization using Cox model-based weighting within the training set (80%). The score weights were fine-tuned via adjustment of cut points for each variable based on clinical interpretability and empirical distributions to optimize predictive performance across time horizons of 1 to 7 years. The final CFRS scoring scheme was recalibrated in the held-out test set (20%) to finalize the point allocation. Discrimination of the final CFRS was evaluated using the C-index and cumulative/dynamic time-dependent AUC. Calibration was assessed by comparing predicted and observed dementia incidence across CFRS-defined risk strata to calculate the integrated calibration index (ICI) and maximum absolute error (Emax). To account for potential overfitting, internal validation was performed using bootstrap optimism correction with 300 resamples^42^ and the optimism-corrected estimates were reported for all performance metrics.

## RESULTS

### FI and Risks of Adverse Clinical Outcomes

The study cohort composed of up to 63,509 dementia-free participants with FI and neuroimaging data (Figure 1). Table 1 summarizes the key characteristics of the cohort. The FI values ranged from 0 to 0.546 (mean = 0.106, SD = 0.065) and exhibited a gamma distribution characteristic of successful aging cohorts^7^ (Supplementary Figure S1a). FI values showed weak correlations with increasing age (r = 0.06, *p* < 0.001) and fewer years of education (r = −0.06, *p* < 0.001) and were slightly higher in females than males (mean difference = 0.011, 95% CI = 0.010–0.012, Welch’s *p* < 0.001). Adjusting for age, sex, and education, higher FI values were associated with increased risks of incident all-cause dementia (HR = 1.55, 95% CI = 1.35–1.78, *p* < 0.001), including AD (HR = 1.43, 95% CI = 1.15– 1.78, *p* = 0.001) and vascular dementia (HR = 1.74, 95% CI = 1.26–2.41, *p* < 0.001) (Supplementary Figure S1b). In addition, higher FI values were associated with increased risks of incident stroke (HR = 1.12, 95% CI = 1.02–1.24, *p* = 0.021) and mortality (HR = 1.27, 95% CI = 1.21–1.34, *p* < 0.001) (Supplementary Figure S1c).

**Table 1.** Cohort characteristics.

| Characteristics | n = 63,509 |
| --- | --- |
| **Demographics** |  |
| Age (mean, SD) | 65.02 (7.68) |
| Sex (n, %) |  |
| Female | 33,243 (52.3) |
| Male | 30,266 (47.7) |
| Ethnicity (White) (n, %) | 61,313 (96.8) |
| Years of education (mean, SD) | 16.62 (4.36) |
| Frailty index (mean, SD) | 0.106 (0.065) |
| **Clinical outcomes (n, %)** |  |
| Incident all-cause dementia<sup>a</sup> | 144 (0.3) |
| Incident Alzheimer's disease dementia<sup>a</sup> | 66 (0.1) |
| Incident vascular dementia<sup>a</sup> | 25 (0.05) |
| Incident stroke<sup>b</sup> | 389 (0.7) |
| Incident mortality<sup>a</sup> | 1,264 (2.3) |
| **Neuroimaging phenotypes (mean, SD)** |  |
| Hippocampal volume (mm<sup>3</sup>)<sup>c</sup> | 7,988.69 (815.85) |
| Global cortical thickness (mm)<sup>c</sup> | 2.66 (0.11) |
| Total white matter hyperintensities volume (mm<sup>3</sup>)<sup>d</sup> | 5,352.56 (7,070.01) |
| Cortical blood flow (mL)<sup>e</sup> | 51.97 (14.37) |
| Global fractional anisotropy<sup>f</sup> | 0.56 (0.02) |
| Global mean diffusivity (mm<sup>2</sup>/s)<sup>f</sup> | 0.00079 (0.00003) |

<sup>a</sup>n = 55,359; <sup>b</sup>n = 54,827; <sup>c</sup>n = 62,336; <sup>d</sup>n = 59,953; <sup>e</sup>n = 5,217; <sup>f</sup>n = 58,991

### FI, Neuroimaging Markers, and their Joint Effects on Clinical Outcomes

Multiple global neuroimaging markers were associated with higher FI values, including, in order of descending effect, higher total WMH volume, lower global FA, lower global CBF, higher global MD, smaller hippocampus, and thinner global cortex (Table 2). After accounting for other neuroimaging markers, total WMH volume (*β* = 0.068, SE = 0.020, *p* < 0.001) and global FA (*β* = −0.082, SE = 0.027, *p* = 0.003) remained significantly associated with higher FI values.

**Table 2.**
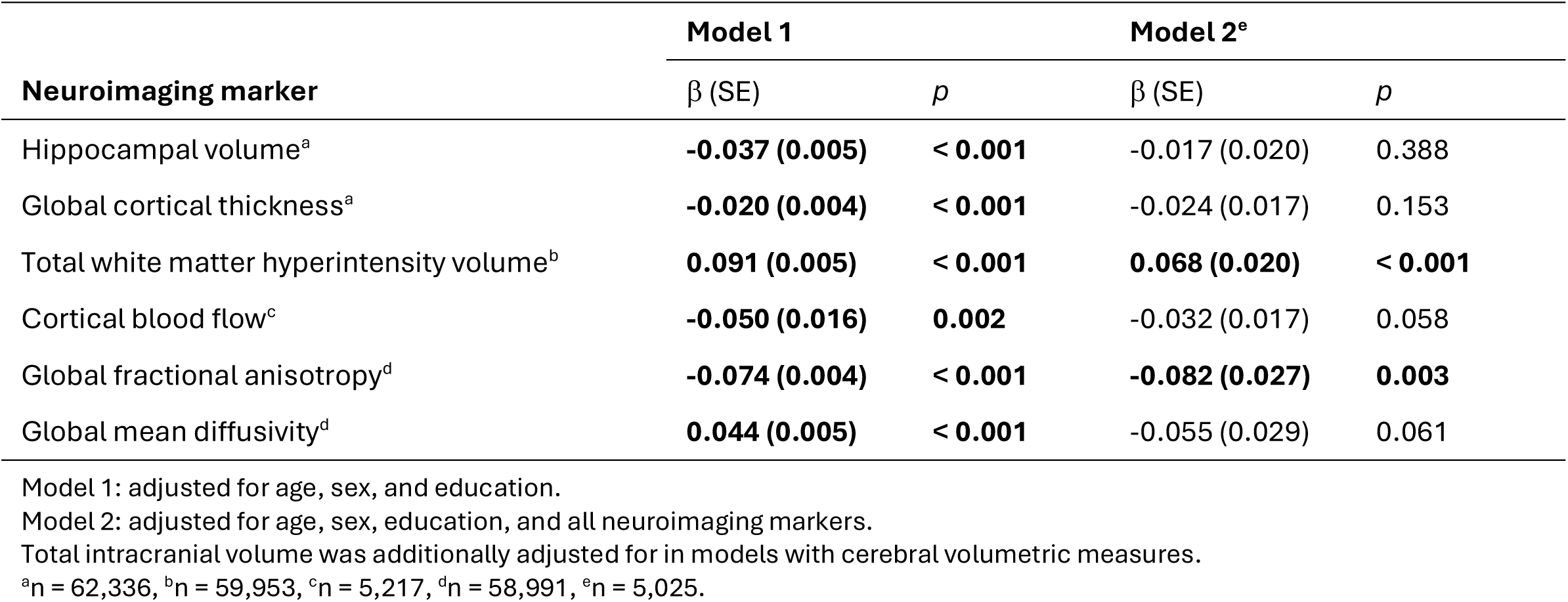
Linear associations of global neuroimaging markers with FI values.

| Neuroimaging marker | Model 1 |  | Model 2<sup>e</sup> |  |
| --- | --- | --- | --- | --- |
| | $\beta$ (SE) | *p* | $\beta$ (SE) | *p* |
| Hippocampal volume<sup>a</sup> | **-0.037 (0.005)** | **< 0.001** | -0.017 (0.020) | 0.388 |
| Global cortical thickness<sup>a</sup> | **-0.020 (0.004)** | **< 0.001** | -0.024 (0.017) | 0.153 |
| Total white matter hyperintensity volume<sup>b</sup> | **0.091 (0.005)** | **< 0.001** | **0.068 (0.020)** | **< 0.001** |
| Cortical blood flow<sup>c</sup> | **-0.050 (0.016)** | **0.002** | -0.032 (0.017) | 0.058 |
| Global fractional anisotropy<sup>d</sup> | **-0.074 (0.004)** | **< 0.001** | **-0.082 (0.027)** | **0.003** |
| Global mean diffusivity<sup>d</sup> | **0.044 (0.005)** | **< 0.001** | -0.055 (0.029) | 0.061 |

Model 1: adjusted for age, sex, and education.
Model 2: adjusted for age, sex, education, and all neuroimaging markers.
Total intracranial volume was additionally adjusted for in models with cerebral volumetric measures.
<sup>a</sup>n = 62,336, <sup>b</sup>n = 59,953, <sup>c</sup>n = 5,217, <sup>d</sup>n = 58,991, <sup>e</sup>n = 5,025.

FI and individual neuroimaging markers were additively associated with increased risks of incident all-cause dementia and mortality, including smaller hippocampus, higher total WMH volume, thinner cortex, higher global MD, and lower global FA (Figure 2, Supplementary Table S2). A significant FI × MD interaction was observed for incident dementia (*β_INT_* = −0.14, HR = 0.87, 95% CI = 0.79–0.96, *p* = 0.004, q = 0.007), where the effect of FI on dementia risk was stronger at lower MD (−1 SD: HR = 1.99, 95% CI = 1.57–2.51) and progressively weakened at higher MD levels (0 SD: HR = 1.73, 95% CI = 1.45–2.07; +1 SD: HR = 1.51, 95% CI = 1.29–1.77). For stroke outcome, higher FI values were associated with increased risk of incident stroke but these effects were attenuated (*p* > 0.05) after accounting for higher total WMH volume (HR = 1.71, 95% CI = 1.52–1.94, *p* < 0.001), higher global MD (HR = 1.36, 95% CI = 1.29–1.44, *p* < 0.001), lower global FA (HR = 1.30, 95% CI = 1.23–1.37, *p* < 0.001), and lower cortical thickness (HR = 1.16, 95% CI = 1.05–1.28, *p* = 0.004) (Supplementary Table S2).

**Figure 2.**
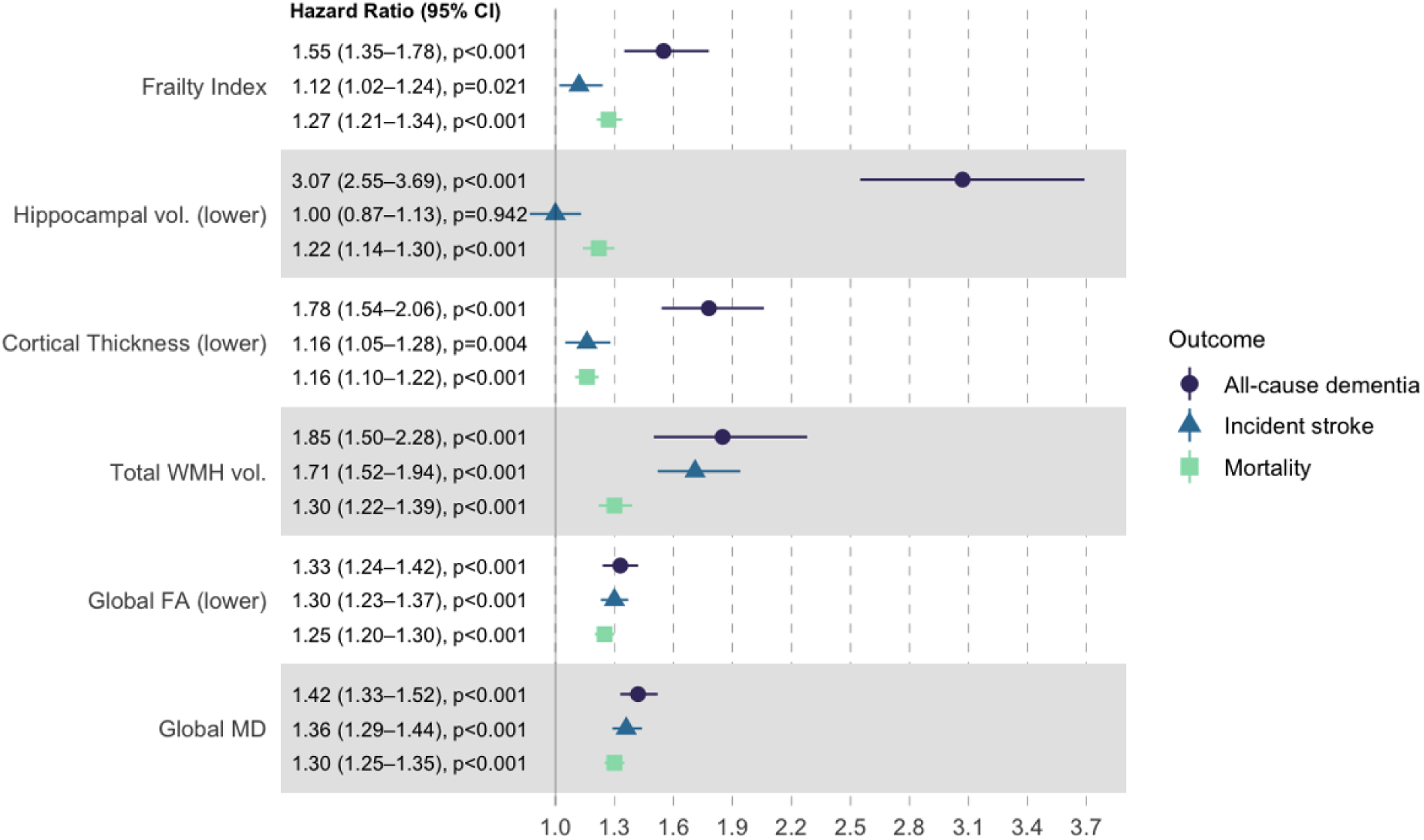
Effects of frailty index and global neuroimaging markers on risks of clinical outcomes. Separate Cox proportional hazards regression models were fitted for each main predictor, adjusted for age, sex, and education. Each neuroimaging model were adjusted for frailty index and models with cerebral volumetric measures were additionally adjusted for total intracranial volume. Abbreviations: CI, confidence interval; FA, fractional anisotropy; MD, mean diffusivity; WMH, white matter hyperintensities.

### Dementia Prediction Model Performance and Interpretation

The cerebral frailty XGBoost-AFT model incorporating neuroimaging phenotypes, FI values, and demographic variables demonstrated strong discrimination (C-index = 0.825, 95% CI = 0.777–0.872) and outperformed models based on demographics and FI values alone (C-index = 0.739, 95% CI = 0.713–0.764) or demographics only (C-index = 0.691, 95% CI = 0.664–0.717). Dementia incidence in the cohort peaked at year 5 and plateaued by year 7. The addition of neuroimaging phenotypes and FI to basic demographic model substantially improved model discrimination and overall prediction accuracy (5-year AUC = 0.854, 95% CI = 0.819–0.890; iBS = 0.019, 95% CI = 0.012–0.023) (Table 3). As illustrated in Figure 3a, the cerebral frailty model maintained consistently stronger and more robust discriminatory capability over most time horizons (iAUC = 0.759, 95% CI = 0.714–0.803), whereas the basic models showed weaker and less stable performance.

**Table 3.**
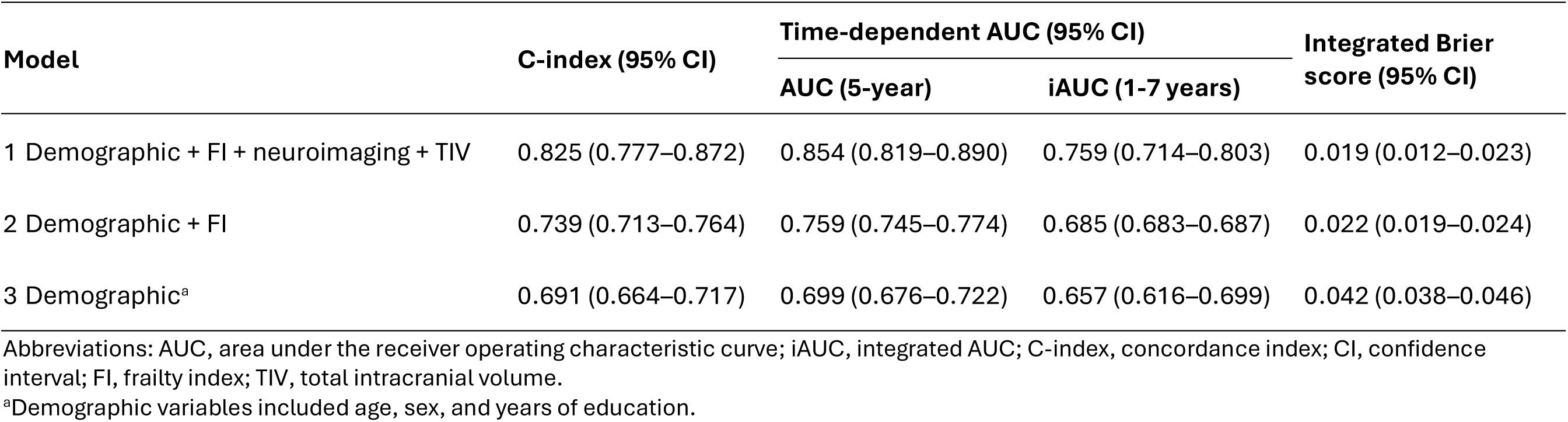
Nested cross-validation performance of XGBoost-Accelerated Failure Time models for incident dementia.

| Model | C-index (95% CI) | Time-dependent AUC (95% CI) |  | Integrated Brier score (95% CI) |
| --- | --- | --- | --- | --- |
|  |  | AUC (5-year) | iAUC (1-7 years) |  |
| 1 Demographic + FI + neuroimaging + TIV | 0.825 (0.777–0.872) | 0.854 (0.819–0.890) | 0.759 (0.714–0.803) | 0.019 (0.012–0.023) |
| 2 Demographic + FI | 0.739 (0.713–0.764) | 0.759 (0.745–0.774) | 0.685 (0.683–0.687) | 0.022 (0.019–0.024) |
| 3 Demographic<sup>a</sup> | 0.691 (0.664–0.717) | 0.699 (0.676–0.722) | 0.657 (0.616–0.699) | 0.042 (0.038–0.046) |

Abbreviations: AUC, area under the receiver operating characteristic curve; iAUC, integrated AUC; C-index, concordance index; CI, confidence interval; FI, frailty index; TIV, total intracranial volume.
<sup>a</sup>Demographic variables included age, sex, and years of education.

**Figure 3.**
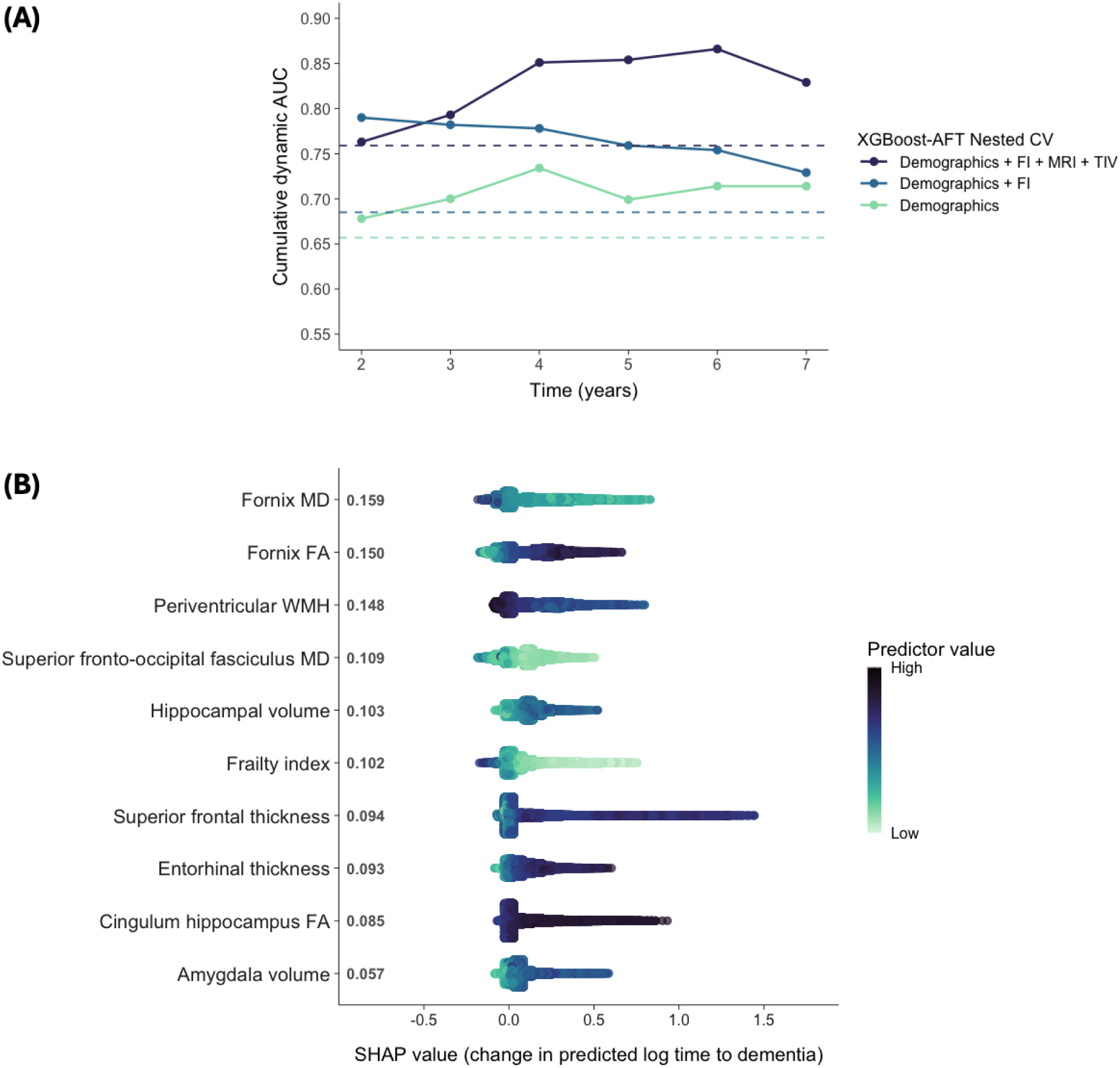
Model discriminatory performance and predictor importance. (A) shows cumulative dynamic AUC across time horizons with integrated AUC (dashed lines) of each model. (B) shows the global SHAP ranking of the 10 most influential variables for model prediction. The position along the x-axis indicates each variable’s impact on time-to-dementia (SHAP < 0 corresponds to shorter time-to-dementia/higher risk) for a given variable magnitude (color bar). Abbreviations: AFT, accelerated failure time; CV, cross-validation; FA, fractional anisotropy; MD, mean diffusivity; SHAP, Shapley Additive exPlanations; TIV, total intracranial volume; WMH, white matter hyperintensities.

Figure 3b presents the mean absolute SHAP value ranking of the 10 most influential variables, where the horizontal position indicates each variable’s impact on predicted time to dementia at a given variable magnitude. Overall, the plot reveals largely monotonic relationships between frailty, neuroimaging phenotypes and dementia risk. Predictors associated with earlier dementia onset included white matter microstructural abnormalities, particularly in the hippocampal circuitry (higher MD and lower FA in the fornix) and the association tract (higher superior fronto-occipital fasciculus MD); greater periventricular WMH burden; higher FI values, and markers of neurodegeneration (thinner superior frontal and entorhinal cortex, and smaller hippocampus and amygdala). In contrast, higher cingulum hippocampus FA was associated with delayed dementia onset.

### Cerebral Frailty Risk Scores and Dementia Prediction

The 10-variable CFRS AutoScore-survival model demonstrated strong discrimination (optimism-corrected C-index = 0.838, 95% CI = 0.794–0.884; 5-year AUC = 0.867, 95% CI = 0.829–0.901; iAUC = 0.778, 95% CI = 0.687–0.845). Table 4 presents the scoring matrix for the dementia prediction model. Variables such as entorhinal thickness, periventricular WMH, and fornix MD received higher point allocation, indicating greater contributions to dementia risk. The total CFRS ranged from 0 to 100 and approximated a normal distribution, with most individuals scoring between 25 and 50 (Figure 4a). Calibration was good across CFRS-defined risk strata, with low absolute miscalibration (optimism-corrected ICI = 0.018, 95% CI = 0.017–0.019) and low error even in extreme risk groups (optimism-corrected Emax = 0.024, 95% CI = 0.022–0.026) (Figure 4b). Increasing CFRS was also associated with progressively earlier dementia onset (log-rank *p* < 0.001), particularly in the top decile (Figure 4c). In a sensitivity analysis using a Fine-Gray regression model treating mortality as a competing event, higher CFRS remained significantly associated with an increased risk of dementia (subdistribution hazard ratio = 1.05, 95% CI = 1.03–1.06, *p* < 0.001).

**Figure 4.**
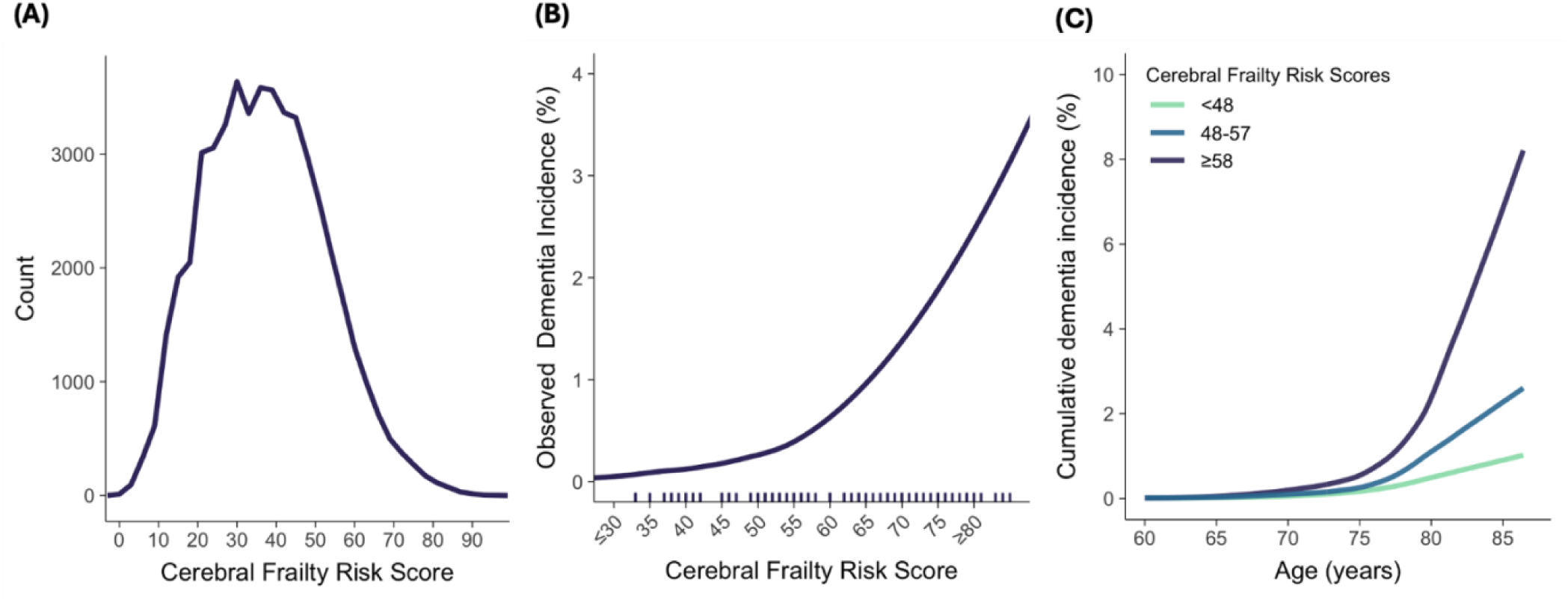
Cerebral frailty risk score (CFRS) derivation and evaluation. (A) shows the CFRS distribution in the UK Biobank. Increasing CFRS were associated with higher dementia incidence (B) and earlier dementia onset (C).

**Table 4.**
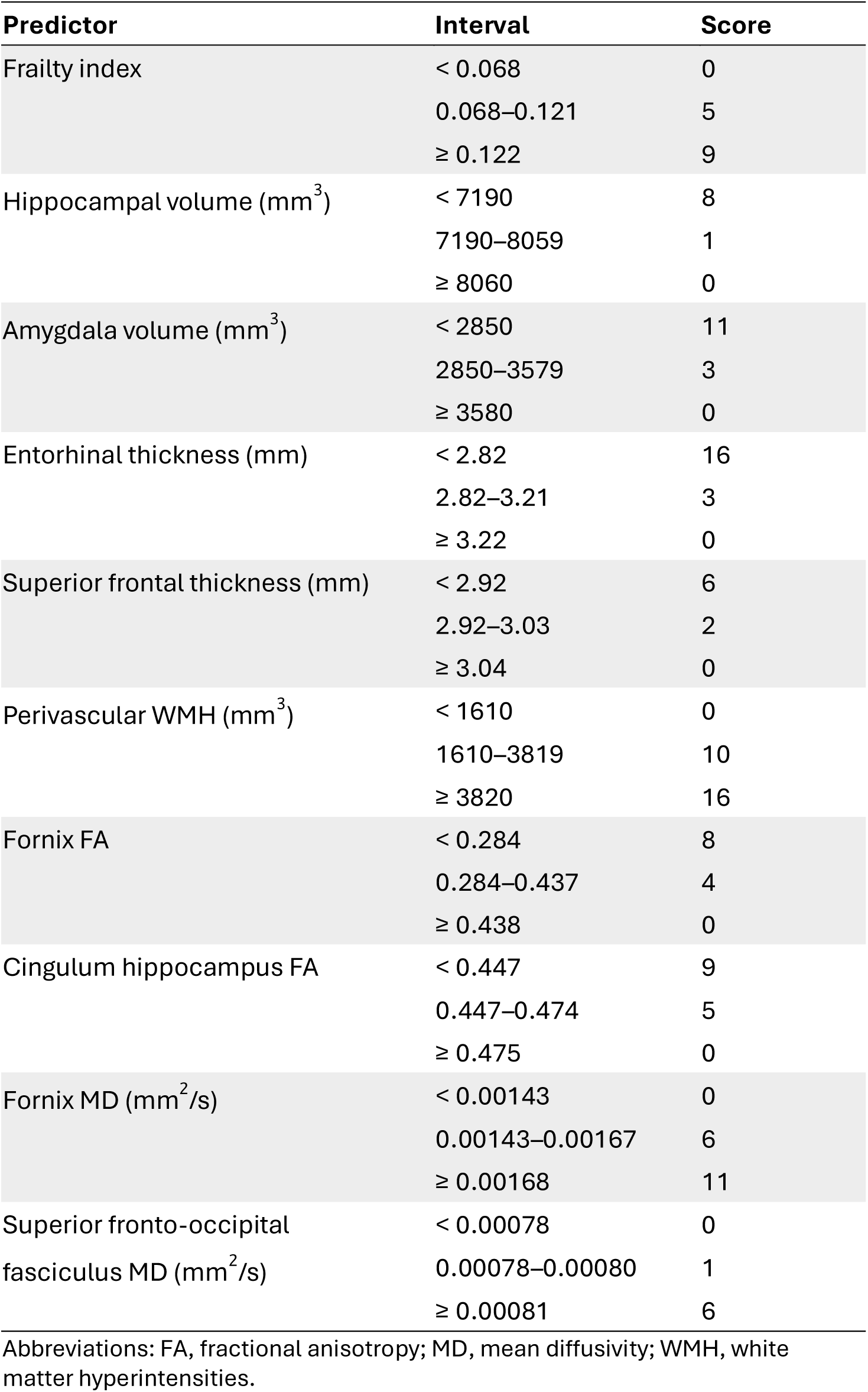
A 10-predictor Cerebral Frailty Risk Score for Dementia.

## DISCUSSION

In the large UK Biobank cohort, higher FI values were associated with multiple indices of brain aging, including greater WMH burden, higher MD, lower FA and CBF, smaller hippocampal volume, and thinner cortex. These features of cerebral dysfunction and FI contributed additively to increased risks of incident all-cause dementia and mortality. The point-based CFRS, which integrated FI and key neuroimaging markers using machine learning demonstrated strong performance in predicting dementia that was robust to competing mortality risk. These findings suggest that frailty reflects cumulative systemic vulnerability and is linked to the accumulation of mixed pathology of cerebrovascular disease, microstructural damage, and neurodegeneration. Declines in both multisystemic and neurophysiological reserves may represent parallel but distinct pathways towards dementia development. The novel CFRS built from frailty and high dimensional neuroimaging features demonstrates potential clinical utility for dementia risk stratification in older populations.

As populations age and individuals live longer with increasing chronic disability and multimorbidity, moving beyond chronological age and disease-centric processes toward frailty-based frameworks may better reflect the substantial heterogeneity in older adults’s health^6^. Frailty as operationalized using the deficit accumulation approach captures a continuum of subclinical to clinical decline across multiple domains of aging. The neuroimaging evidence on FI is sparse and confined to memory clinic cohorts that included dementia patients. These studies yielded inconsistent results with one associating higher FI with the presence of cortical atrophy^19^, while the other found no relationship with neurodegeneration markers but linked FI to markers of cerebral small vessel disease^17^. Among studies examining the Fried phenotype, findings on WMH volume have been inconsistent^13,15,16,18^, but several have linked frailty or its specific components (weakness/slowness) to cerebral atrophy^13,16,18,20^, cortical infarcts^18^, and markers of white matter integrity^15^.

We observed that higher FI was associated with mixed neurodegenerative and vascular pathology, with comparatively stronger associations for cerebrovascular (higher WMH volume, lower CBF) and microstructural (lower FA, higher MD) dysfunction. This suggests that frailty reflects a broader multisystem process, wherein age-related microvascular changes and possible AD-related pathology may play a central role in its pathophysiology. Consistent with this interpretation, individuals with higher FI showed increased risks of incident stroke, all-cause dementia, and mortality. However, the relative contribution of FI to incident stroke risk was smaller and not statistically significant when compared to neuroimaging markers of brain health, particularly WMH burden. Although the FI already incorporates vascular risk factors and cardiometabolic history, elevated FI may itself reflect a clinical manifestation of underlying cerebrovascular pathology. Conversely, given the heterogeneity of stroke in terms of extent, location, and subtype, future studies may examine how stroke severity and outcomes may be influenced by pre-stroke frailty^43^ and brain or heart health.

In predicting incident dementia, we observed a negative interaction between FI and MD, such that the adverse effect of frailty on dementia risk was strongest at lower diffusivity values (preserved white matter integrity) and attenuated at higher diffusivity values (impaired white matter integrity). MD derived from DTI quantifies the degree of water molecules diffusion and is sensitive to subtle white matter microstructural changes in the aging brain^44^. This pattern suggests that additional systemic vulnerability may confer comparatively less incremental risk in the presence of substantial microstructural damage, but may exert greater influence on dementia risk when brain microstructural reserve remains relatively intact. Accordingly, frailty assessments that capture the composite burden of clinical deficits and functional impairment may be particularly valuable for identifying individuals at elevated risk of cognitive decline during early stages of white matter pathology. Moreover, as frailty remained associated with dementia risk independent of neuroimaging markers, this also suggests that frailty intervention may represent a promising strategy for dementia prevention. The additive effects of frailty and brain dysfunction capturing cerebrovascular, neurodegenerative, and microvascular burden on dementia risk add to the evidence that dementia is a multifactorial condition arising from increasing cerebral pathological burden alongside decreasing resilience against the accumulation of age-related health deficits.

When further examining the clinical relevance of FI in conjunction with large-scale neuroimaging phenotypes, the machine learning approach demonstrated strong predictive performance for dementia risk, outperforming more basic models despite low prevalence in the cohort. The resulting cerebral frailty model identified higher FI and multiple regions-of-interest reflecting mixed brain pathology as key contributors to earlier dementia onset. Notably, medial temporal lobe dysfunction emerged as a critical predictor, at both the structural level (reduced hippocampal and amygdala volumes and thinner entorhinal cortex) and network level (higher MD and lower FA in the fornix, as well as higher FA in the hippocampal cingulum as a protective factor). Alterations in the frontal region were also prominent, including higher MD in the superior fronto-occipital fasciculus and thinner superior frontal cortex. These frontal and temporal regions are well-established in supporting cognitive function and are known to be particularly vulnerable in neurodegenerative processes^45–47^. In addition, a greater periventricular WMH burden emerged as a critical cerebrovascular contributor to accelerated dementia onset, potentially attributable to underlying endothelial dysfunction, interstitial edema, and impaired glymphatic clearance pathway^48^. The current work extend prior autopsy study demonstrating a cumulative association between FI with mixed neuropathologies (e.g. β-amyloid, α-synuclein, arteriolosclerosis, cerebral infarcts) and dementia diagnosis^49^, reinforcing the integrative role of systemic frailty and cerebral pathological burden in the mechanistic pathways of dementia.

Although the overall predictive performance of the FI-based model that combined FI, age, sex, and education was modest, the inclusion of FI outperformed the parsimonious model based on demographic factors only, underscoring frailty as an important systemic risk factor and preventive target for dementia. By integrating key indices of frailty and brain health into an interpretable scoring framework, the CFRS offers a potential enrichment tool for dementia prevention trials and risk stratification, with implications for early detection, patient care, and healthcare resource allocation. While the current use of high-dimensional neuroimaging data may limit widespread clinical implementation, the CFRS introduces a novel concept that captures the cumulating deficits of cerebral frailty for detecting pathological cognitive decline beyond normal aging. Importantly, population-specific FI measure could be generated, given the robustness of FI to the number and selection of deficit items^3^ and the consistency of FI-related epidemiological findings across geographical regions and diverse demographic groups^50,51^. In parallel, advances in neuroimaging, such as deep learning techniques for shorter scan time^52,53^, and increased clinical adoption of automated volumetric and lesion segmentation pipelines^54,55^ are lowering barriers to clinical translation. As quantitative neuroimaging and automatic processing become more accessible, data-driven tools such as the CFRS could potentially be incorporated into routine clinical practice to enable more sensitive risk profiling, support personalized interventions, and improve patient outcomes.

One of the strengths of this study is the use of a large prospective population-based dataset with a validated frailty measure. The UK Biobank’s rich integration of clinical, imaging, and genetic data provides opportunities for future investigations of FI with additional domains (e.g. retinal or cardiac imaging^56^), which may contribute further insights into the etiology of frailty. Methodologically, this study combined predictive modelling (XGBoost-AFT) with explanatory modelling (Autoscore-Survival), which provided a structured way to translate complex survival models into clinically intepretable risk scores. XGBoost-AFT retains the advantages of tree-based models (e.g efficient analysis of large-scale data, tolerates multicollinearity and mixed data types, captures nonlinear associations) while natively accommodating right-censoring and enabling risk stratification across time horizons, making it well suited for time-sensitive and age-dependent diseases^22,24,57^. Integrating Autoscore further facilitated the identification of key risk factors while maintaining a balance between predictive performance, model complexity, and interpretability of diverse biomedical data^23,58^. Such an integrative machine learning approach may be valuable for future analyses of large clinical or registry datasets involving survival outcomes.

However, our results should be interpreted with caution due to several limitations. First, the UK Biobank cohort is subject to a degree of a “healthy volunteer” bias^5^, particularly among participants who completed neuroimaging as they were younger and had slightly lower FI values than those without imaging. The prevalence of incident dementia in this cohort was lower than in other cohorts, which may have contributed to the wider confidence intervals and an increased risk of model overfitting. Although we have implemented several mitigation measures, including internal validation approaches suited for low-event samples^35,42^, statistical adjustment for class imbalance, and model regularization, performance estimates may still show high variance and instability given the limited number of events. External validation in independent cohorts is therefore necessary to confirm the robustness and generalizability of our findings. Second, our analyses were based on cross-sectional FI and neuroimaging data, which precluded causal inferences regarding the temporal relationship between frailty and brain health. Nevertheless, the UK Biobank has a sizable subset of participants returning for repeat assessments, providing an opportunity for future longitudinal investigations. Third, no single machine learning algorithm is universally optimal and model performance may vary across populations and clinical contexts. Hence, direct comparisons with existing dementia prediction models could be challenging due to differences in cohort characteristics, outcome definitions, and feature sets.

In conclusion, frailty reflects systemic vulnerability and is associated with mixed cerebral pathology of cerebrovascular disease, microstructural damage, and neurodegeneration. Both systemic and neural factors may represent parallel yet distinct pathways to dementia. By integrating frailty and cerebral dysfunction, the CFRS methodology demonstrates the potential of applying a machine learning survival analysis approach to large-scale population-based data to enhance dementia risk stratification and guiding preventive interventions to improve clinical outcomes.

## Data Availability

All data produced in the present study are available upon reasonable request to the authors.

## ACKNOWLEDGEMENTS

This research has been conducted using the UK Biobank Resource under Application Number 243130. This study was supported by MOE Tier 1 RG58/23 and the Social Science Research Council (Singapore), administered by the Ministry of Education, Singapore, under its Social Science and Humanities Research (SSHR) Fellowship (SSRC2023-SSHR-003).

## ETHICS DECLARATIONS

The authors declare no competing interests.

## SUPPLEMENTARY TABLES

**Table S1.** List of variables used for XGBoost-AFT model training.

|  |  |  |
| --- | --- | --- |
| cth_banks_of_the_superior_temporal_sulcus | cth_caudal_anterior_cingulate | cth_caudal_middle_frontal |
| cth_cuneus | cth_entorhinal | cth_frontal_pole |
| cth_fusiform | cth_inferior_parietal | cth_inferior_temporal |
| cth_insula | cth_isthmus_cingulate | cth_lateral_occipital |
| cth_lateral_orbitofrontal | cth_lingual | cth_medial_orbitofrontal |
| cth_middle_temporal | cth_paracentral | cth parahippocampal |
| cth_pars_opercularis | cth_pars_orbitalis | cth_pars_triangularis |
| cth_pericalcarine | cth_postcentral | cth_posterior_cingulate |
| cth_precentral | cth_precuneus | cth_rostral_anterior_cingulate |
| cth_rostral_middle_frontal | cth_superior_frontal | cth_superior_parietal |
| cth_superior_temporal | cth_supramarginal | cth_transverse_temporal |
| total_intracranial_volume | vol_bankssts | vol_caudalanteriorcingulate |
| vol_caudalmiddlefrontal | vol_cuneus | vol_entorhinal |
| vol_frontalpole | vol_fusiform | vol_inferiorparietal |
| vol_inferiortemporal | vol_insula | vol_isthmuscingulate |
| vol_lateraloccipital | vol_lateralorbitofrontal | vol_lingual |
| vol_medialorbitofrontal | vol_middletemporal | vol_paracentral |
| vol parahippocampal | vol_parsopercularis | vol_parsorbitalis |
| vol_parstriangularis | vol_pericalcarine | vol_postcentral |
| vol_posteriorcingulate | vol_precentral | vol_precuneus |
| vol_rostralanteriorcingulate | vol_rostralmiddlefrontal | vol_superiorfrontal |
| vol_superiorparietal | vol_superiortemporal | vol_supramarginal |
| vol_transversetemporal | vol_accumbens | vol_amygdala |
| vol_caudate | vol_hippocampus | vol_pallidum |
| vol_putamen | vol_thalamus | fa_anteriorcoronaradiata |
| fa_anteriorlimbofinternalcapsule | fa_bodyofcorpuscallosum | fa_cerebralpeduncle |
| fa_cingulumcingulategyrus | fa_cingulumhippocampus | fa_corticospinaltract |
| fa_externalcapsule | fa_fornixcres_striaterminalis | fa_fornix |
| fa_genuofcorpuscallosum | fa_inferiorcerebellarpeduncle | fa_mediallemniscus |
| fa_middlecerebellarpeduncle | fa_pontinecrossingtract | fa_posteriorcoronaradiata |
| fa_posteriorlimbofinternalcapsule | fa_posteriorthalamicradiation | fa_retrolenticularpartofinternalcapsule |
| fa_sagittalstratum | fa_spleniumofcorpuscallosum | fa_superiorcerebellarpeduncle |
| fa_superiorcoronaradiata | fa_superiorfrontooccipitalfasciculus | fa_superiorlongitudinalfasciculus |
| fa_tapetum | fa_uncinatefasciculus | md_anteriorcoronaradiata |
| md_anteriorlimbofinternalcapsule | md_bodyofcorpuscallosum | md_cerebralpeduncle |
| md_cingulumcingulategyrus | md_cingulumhippocampus | md_corticospinaltract |
| md_externalcapsule | md_fornixcres_striaterminalis | md_fornix |
| md_genuofcorpuscallosum | md_inferiorcerebellarpeduncle | md_mediallemniscus |
| md_middlecerebellarpeduncle | md_pontinecrossingtract | md_posteriorcoronaradiata |
| md_posteriorlimbofinternalcapsule | md_posteriorthalamicradiation | md_retrolenticularpartofinternalcapsule |
| md_sagittalstratum | md_spleniumofcorpuscallosum | md_superiorcerebellarpeduncle |
| md_superiorcoronaradiata | md_superiorfrontooccipitalfasciculus | md_superiorlongitudinalfasciculus |
| md_tapetum | md_uncinatefasciculus | deep_wmh |
| periventricular_wmh | age | education_years |
| Sex.Female | Sex.Male | FI_value |

**Table S2.** Cox proportional hazards regression predicting clinical outcomes.

| Model | All-cause dementia |  | All-cause mortality |  | Incident stroke |  |
| --- | --- | --- | --- | --- | --- | --- |
|  | HR (95% CI) | P | HR (95% CI) | P | HR (95% CI) | P |
| 1 Frailty index | **1.55 (1.35, 1.78)** | **8.27e-10** | **1.27 (1.21, 1.34)** | **<2e-16** | **1.12 (1.02, 1.24)** | **0.021** |
| 2 Hippocampal volume (lower) | **3.07 (2.55, 3.69)** | **< 2e-16\*** | **1.22 (1.14, 1.30)** | **1.6e-09\*** | 1.00 (0.87, 1.13) | 0.942 |
| Frailty index | **1.55 (1.35, 1.80)** | **1.97e-09†** | **1.25 (1.19, 1.32)** | **< 2e-16†** | 1.12 (1.01, 1.24) | 0.026 |
| Hippocampal*Frailty | 0.97 (0.84, 1.13) | 0.707 | 1.04 (0.99, 1.10) | 0.119 | 0.98 (0.89, 1.08) | 0.688 |
| 3 Cortical thickness (lower) | **1.78 (1.54, 2.06)** | **3.16e-15\*** | **1.16 (1.10, 1.22)** | **4.56e-08\*** | **1.16 (1.05, 1.28)** | **0.003\*** |
| Frailty index | **1.55 (1.35, 1.79)** | **9.73e-10†** | **1.26 (1.19, 1.32)** | **< 2e-16†** | 1.11 (1.01, 1.23) | 0.033 |
| Cortical thickness*Frailty | 0.88 (0.79, 0.99) | 0.031 | 1.05 (1.00, 1.10) | 0.054 | 1.04 (0.95, 1.14) | 0.355 |
| 4 Total WMH | **1.85 (1.50, 2.28)** | **8.08e-09\*** | **1.30 (1.22, 1.39)** | **< 2e-16\*** | **1.71 (1.52, 1.94)** | **< 2e-16\*** |
| Frailty index | **1.49 (1.27, 1.74)** | **6.01e-07†** | **1.24 (1.17, 1.31)** | **1.10e-14†** | 1.09 (0.98, 1.21) | 0.113 |
| WMH*Frailty | 0.89 (0.76, 1.05) | 0.165 | 1.05 (0.99, 1.10) | 0.108 | 1.04 (0.93, 1.15) | 0.510 |
| 5 Global FA (lower) | **1.33 (1.24, 1.42)** | **1.70e-15\*** | **1.25 (1.20, 1.30)** | **< 2e-16\*** | **1.30 (1.23, 1.37)** | **< 2e-16\*** |
| Frailty index | **1.54 (1.31, 1.81)** | **1.88e-07†** | **1.24 (1.17, 1.31)** | **4.5e-14†** | 1.09 (0.98, 1.21) | 0.126 |
| FA*Frailty | 0.93 (0.85, 1.02) | 0.124 | 1.00 (0.95, 1.04) | 0.850 | 1.00 (0.93, 1.06) | 0.898 |
| 6 Global MD | **1.42 (1.33, 1.52)** | **< 2e-16\*** | **1.30 (1.25, 1.35)** | **< 2e-16\*** | **1.36 (1.29, 1.44)** | **< 2e-16\*** |
| Frailty index | **1.54 (1.31, 1.82)** | **1.75e-07†** | **1.24 (1.17, 1.31)** | **3.44e-14†** | 1.09 (0.98, 1.21) | 0.130 |
| MD*Frailty | **0.87 (0.79, 0.96)** | **0.004\*** | 1.00 (0.96, 1.04) | 0.897 | 0.95 (0.89, 1.02) | 0.172 |

Adjusted for age, sex, education. Separate models were fitted to test for interaction between each MRI marker and frailty index. Total intracranial volume was additionally adjusted in models with cerebral volumetric measures.
\*q < 0.05 (10 comparisons); †q < 0.05 (5 comparisons).
Abbreviations: CI, confidence interval; FA, fractional anisotropy; MD, mean diffusivity; WMH, white matter hyperintensities.

## SUPPLEMENTARY FIGURES

**Figure S1.**
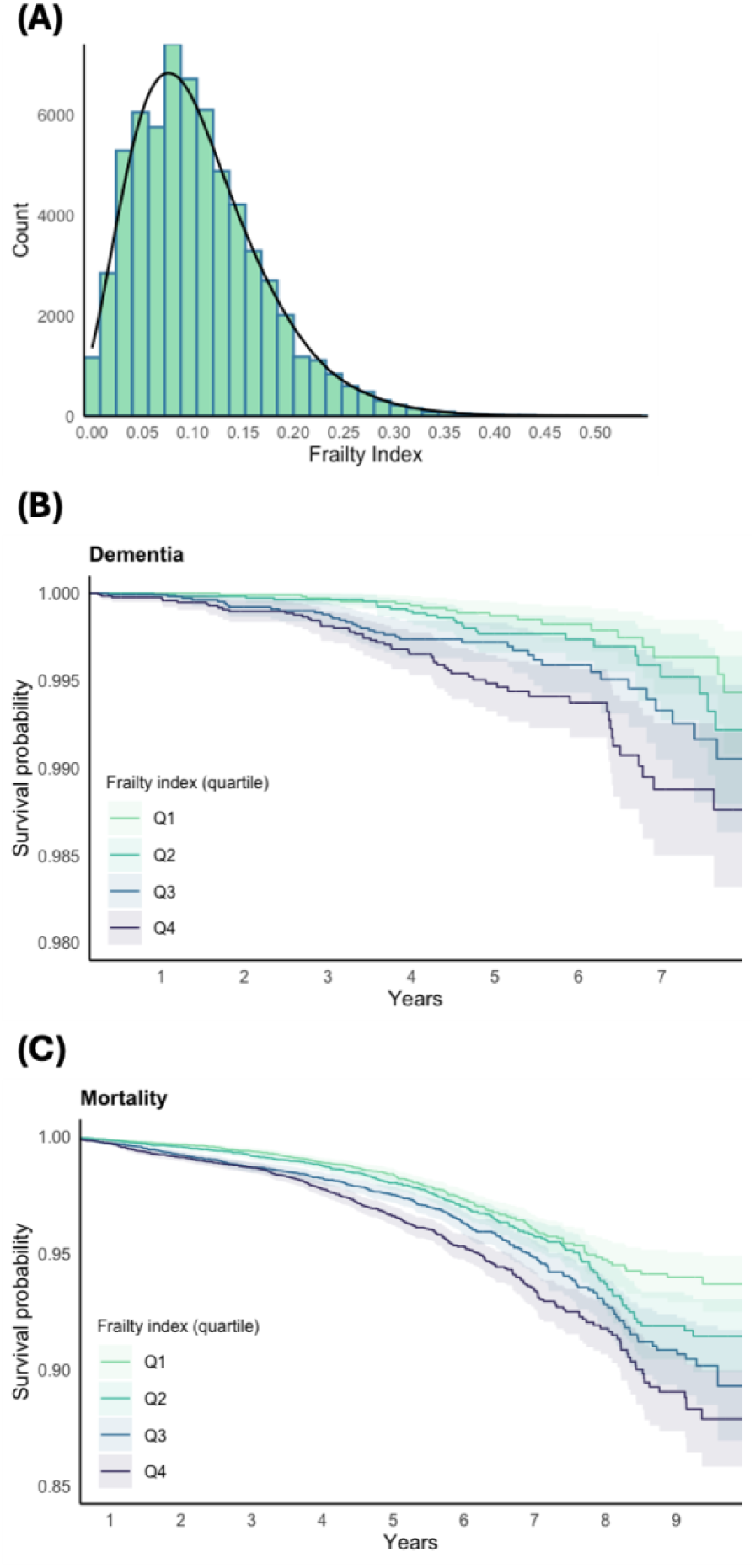
Distribution and survival curves of FI values. The FI values showed a gamma distribution in the UK Biobank cohort (A). Higher FI values were associated with increased risks of incident all-cause dementia (B) and mortality (C), adjusted for age, sex, and education.

